# Trends in AI-powered classification of thyroid neoplasms based on histopathology images, a systematic review

**DOI:** 10.1101/2023.09.24.23295995

**Authors:** Haitham Kussaibi, Noor Alsafwani

**Affiliations:** Department of Pathology, college of Medicine, Imam Abdulrahman bin Faisal University, P.O. Box 1982, Dammam 31441, Saudi Arabia

**Keywords:** Thyroid neoplasms, histopathology images, artificial intelligence, machine learning, deep learning, computer-aided/assisted classification

## Abstract

**Background:** Assessment of thyroid nodules histopathology using AI is crucial for an accurate diagnosis. This systematic review analyzes recent works employing deep learning approaches for classifying thyroid nodules based on histopathology images, evaluating their performance, and identifying limitations.

**Methods:** Eligibility criteria focused on peer-reviewed English papers published in the last five years, applying deep learning to categorize thyroid histopathology images. The PubMed database was searched using relevant keyword combinations.

**Results:** Out of 103 articles, 11 studies met inclusion criteria. They used convolutional neural networks to classify thyroid neoplasm. Most studies aimed at basic tumor subtyping; however, three studies targeted the prediction of tumor-associated mutation. Accuracy ranged from 77% to 100%, with most over 90%.

**Discussion:** The findings from our analysis reveal the effectiveness of deep learning in identifying discriminative morphological patterns from histopathology images, thus enhancing the accuracy of thyroid nodule histopathological classification. Key limitations were small sample sizes, subjective annotation, and limited dataset diversity. Further research with larger diverse datasets, model optimization, and real-world validation is essential to translate these tools into clinical practice.

**Other:** *Funding:* This study did not receive any funding.

*Registration:* The procedural instructions for this systematic review were officially recorded within the PROSPERO database under registration number **RD42023457854** https://www.crd.york.ac.uk/Prospero/

## 1. Introduction

Thyroid nodules are frequently observed in clinical practice, and accurate diagnosis is crucial for proper patient management. The accurate characterization and classification of thyroid tumors, including distinguishing malignant from benign lesions, remains a significant challenge in pathology. Several diagnostic modalities, including radiology US (ultrasound), CT (computerized tomography) scan, MRI (magnetic resonance imaging), cytopathology, and histopathology, are in place to achieve this goal. While histopathology examination is considered the gold standard for diagnosis, it can be sometimes challenging due to the subjective nature of interpretation by pathologists. Manual evaluation of thyroid histopathology slides is prone to interobserver variability, especially in borderline lesions such as NIFTP (Noninvasive Follicular Thyroid Neoplasm with Papillary-like Nuclear Features) (1).

A common theme is developing computational approaches to complement pathologists’ expertise and reduce the subjectivity and interobserver variability inherent to manual slide review and conventional microscopy. The utilization of AI (artificial intelligence) methods has demonstrated the potential to enhance the precision and efficiency of thyroid nodule histopathology diagnosis. Emerging DL (deep learning) techniques offer the potential to improve the efficiency and consistency of analyzing complex morphological patterns in thyroid histologic images.

Recent studies have investigated the utility of deep CNNs (convolutional neural networks) and related DL approaches for the computational pathology of thyroid lesions (2, 3). They have focused specifically on developing DL systems to classify thyroid tumors/nodules into different subtypes including malignant vs. benign categories to guide clinical decision-making.

Other than DL, some studies adopted a traditional image analysis approach for thyroid nodule classification based on nuclear morphology characterization (4, 5). **Valentim** et al. used computerized nuclear morphometry and texture analysis of histology images coupled with CRT (Classification and Regression Trees) to distinguish follicular pattern thyroid lesions. They demonstrated that computerized quantitative image analysis can also provide discriminating nuclear features to classify thyroid tumors (6). While accurate for the study dataset, extensive feature engineering and limited diversity may affect scalability.

Although DL-assisted classification of thyroid nodules based on radiology (7) or cytopathology (8-10) images are extensively explored in the literature, no recent review evaluated the performance of such techniques in classifying histopathology images.

The application of digital pathology and DL for the analysis of thyroid histopathology images is an emerging area of research. While the approaches and aims vary, the collective work highlights the significant potential of these techniques to uncover novel insights from high-resolution WSIs (whole slide images).

Herein, we aim, in this systematic review, to identify and summarize recent studies (published within the past 5 years) employing DL approaches in classifying thyroid neoplasms based on histopathology images. The research’s primary objectives are to evaluate these methods’ effectiveness, identify prevailing trends and patterns, and pinpoint areas that require further investigation.

## 2. Method

### 2.1 Search Strategy

We conducted this systematic review following the PRISMA (Preferred Reporting Items for Systematic Reviews and Meta-Analysis) guidelines (11). The review protocol was registered within the PROSPERO database under registration number CRD42023457854.

We performed a comprehensive literature search of the Medline PubMed database, focusing on the last 5 years from January 2019 to September 2023. We utilized specific combinations of keywords such as ‘thyroid’, ‘histopathology’, ‘images’, ‘classification’, ‘diagnosis’, ‘prediction’, ‘artificial intelligence’, ‘machine learning’, ‘deep learning’, ‘computer-aided/assisted’, and other relevant terms. Our search was limited to English language original research articles published in high-quality, peer-reviewed journals indexed in PubMed. To ensure inclusion of the most relevant research, we iteratively refined the PubMed search query using various keyword combinations. The final optimized search query was:

(Thyroid[Title] AND (Histopatholog*[Tiab] OR Histolog*[Tiab] OR Patholog*[Tiab]) AND (image*[Tiab] OR slide*[Tiab]) AND (classif*[Tiab] OR diagnos*[Tiab] OR predict*[Tiab]) AND (artificial intelligence[Tiab] OR machine learning[Tiab] OR deep learning[Tiab] OR convolutional[Tiab] OR neural network*[Tiab] OR automated[Tiab] OR computer-assisted[Tiab] OR computer-aided[Tiab]))

### 2.2 Study Selection

The first step in refining the search results was screening the titles and abstracts to identify relevant works based on predetermined inclusion and exclusion criteria.

The inclusion criteria were as follows: 1) original English-language research articles published within the last 5 years and indexed in PubMed that 2) utilized AI/ML/DL techniques for classification of thyroid neoplasms based on histopathology images, 3) reported performance metrics of the classification methods, and 4) had full text freely available.

The exclusion criteria were: 1) studies based on cytology images, 2) studies based on radiology images (US, CT, MRI, etc.), and 3) studies focused solely on morphometry and/or feature engineering.

### 2.3 Data Extraction

We extracted pertinent information from the studies selected during the screening process using a standardized form. The extracted data included details such as author, title, objectives, data characteristics (source, type, quantity), study design (methodological approach), ML/DL model architecture including input patch dimensions and output classes, evaluation metrics, and limitations as reported by the authors. We performed a descriptive analysis of the extracted data to identify prevalent patterns and trends.

## 3. Results

### 3.1 Overview of the included studies (Fig. 1)

The search strategy outlined above initially yielded 103 articles. After excluding non-English and non-original articles, 95 articles remained. Screening the titles and abstracts based on the predefined inclusion and exclusion criteria led to the exclusion of papers utilizing imaging modalities other than histopathology (e.g. cytopathology, ultrasound, CT, MRI). This screening process resulted in 15 remaining articles, of which 11 had freely accessible full text.

**Figure 1:**
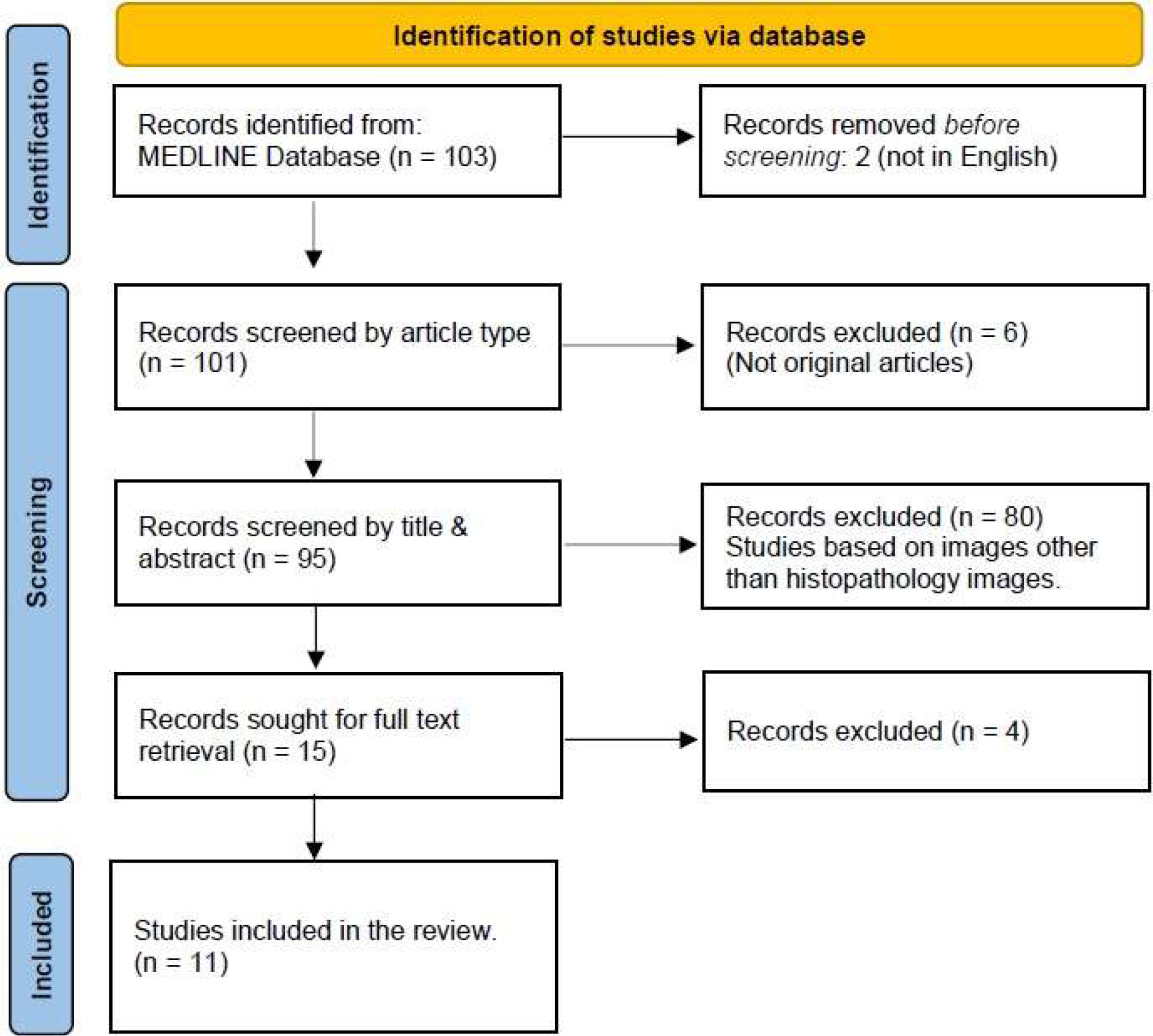
Prisma Flow Diagram

### 3.2 Study Characteristics

A total of 11 studies meeting the inclusion criteria were identified. These studies were published between January 2019 and August 2023. The key trends observed across the included studies were: 1) leveraging whole slide images from histopathology glass slides; 2) extracting image patches/tiles to generate large, labeled datasets suitable for deep learning; 3) utilizing state-of-the-art CNNs pretrained on natural images (ImageNet) and fine-tuned on histology data; 4) incorporating image preprocessing and augmentation strategies to improve model performance; 5) complementing deep learning with traditional image analysis and machine learning where limited labeled data was available; 6) focusing primarily on basic classification of thyroid neoplasms; 7) demonstrating promise in predicting specific genetic mutations linked to malignant behavior and prognosis from histomorphology patterns; 8) experimenting with training CNN models on hyperspectral rather than traditional RGB images; and 9) utilizing frozen rather than formalin-fixed paraffin-embedded sections in 3 studies.

In addition to the 11 included studies, several other relevant studies with similar objectives were identified but excluded due to unavailable full text access. These included a study by Anand et al. 2021 (12) that described a weakly supervised technique to train a DNN (deep neural network) to predict BRAF V600E mutational status with high accuracy in unannotated H&E-stained images of thyroid cancer tissue. Two studies by Esce et al. ((13, 14)) demonstrated the efficacy of CNNs in predicting the probability of nodal metastases in PTC (papillary thyroid carcinoma) based on the histopathology features of the primary tumor. Finally, a study by Nojima et al. 2023 (15) compared the prediction accuracy of 3 CNN models in distinguishing FA (follicular adenoma) from FTC (follicular thyroid carcinoma) based on histopathology images.

## 4. Discussion

This systematic review demonstrates current applications of deep learning and digital pathology techniques for the automated analysis of thyroid histopathology images. The collection of included studies highlights a diversity of approaches with the shared goal of leveraging digital pathology imaging and artificial intelligence to enhance the analysis of thyroid histopathology specimens. Although all studies utilized histopathological tissue images as the dataset, there were some key differences in their approaches. The studies differed in terms of specific objectives, sample types, image formats, and analytical methodology. Nevertheless, the results collectively demonstrated the utility of machine learning and deep learning-based techniques for enabling enhanced classification and diagnosis based on thyroid histopathology images.

### 4.1 Comparing Objectives: Exploring Simple Subtyping vs. Predicting Associated Mutations

The specific classification tasks and objectives varied across the included studies. Although the principal aim for most studies was basic diagnostic classification of thyroid neoplasms with generic tumor subtyping, which has direct diagnostic relevance, three studies focused on the novel goal of predicting specific genetic mutations associated with thyroid tumors directly from histomorphology features (16-18). These studies targeted mutations such as TERT, BRAF, and RAS, demonstrating the potential to infer molecular characteristics from histological imaging features (table1).

**Table 1:**
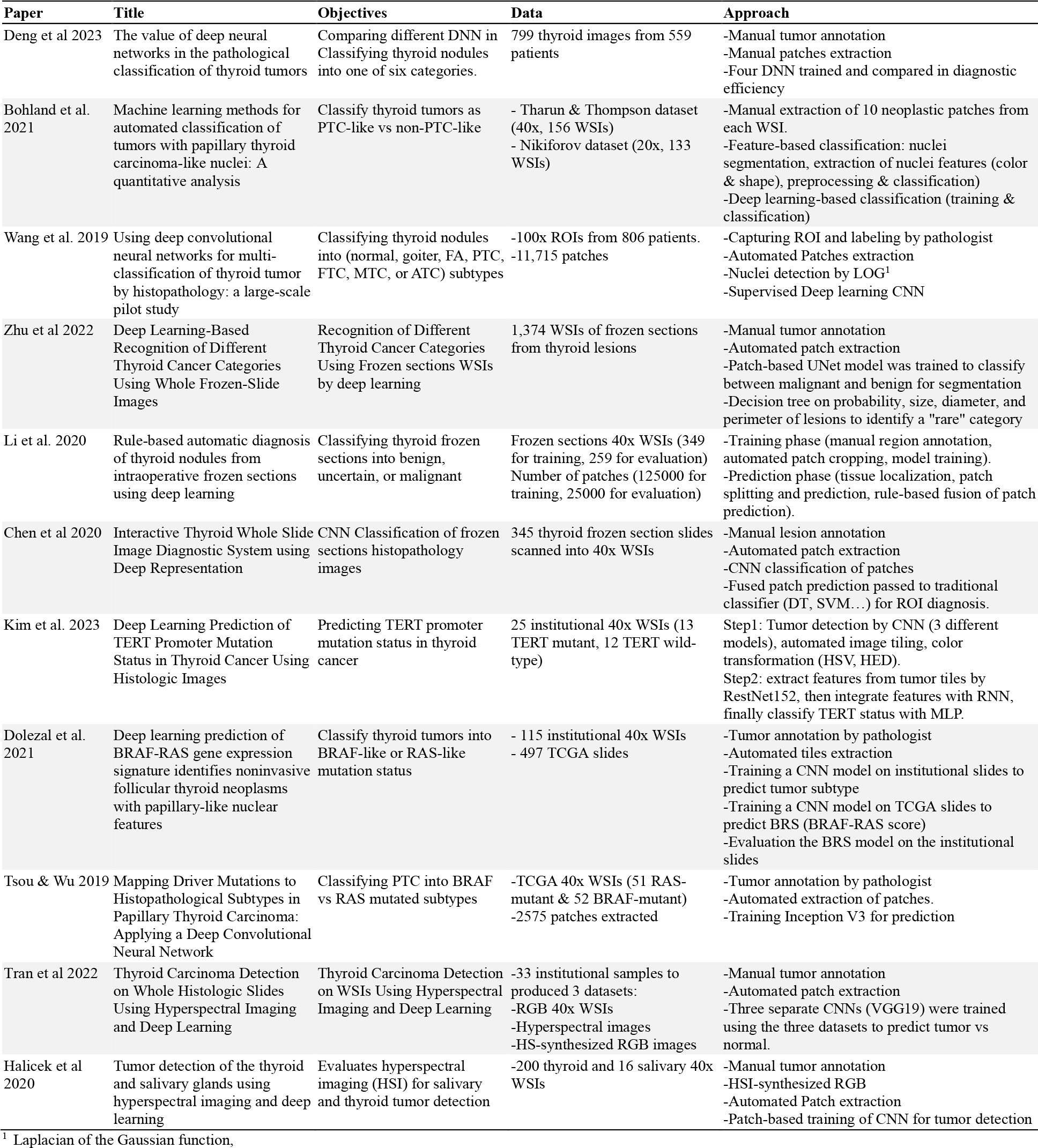
overview of the selected studies including their objectives, methodologies and data used.

### 4.2 Tissue sample preparation, processing, and staining

The sample sources and preparation methods varied among the included studies. While most utilized formalin-fixed paraffin-embedded (FFPE) thyroid tissue sections, three studies (19-21) used frozen sections. All samples were stained with the conventional hematoxylin and eosin (H&E) protocol.

Although H&E staining is the histopathology standard, Ozolek et al. 2014 (5) explored special stains such as Feulgen stain, which preferentially binds DNA and highlights nuclear chromatin patterns better than H&E. Enhanced visualization of nuclear features is particularly relevant in thyroid tumors, where nuclear characteristics are key for precise classification. While H&E remains the typical staining technique, evaluations of complementary stains like Feulgen may further improve the nuclear detail essential for computational pathology applications in this domain (5).

### 4.3 Data sources: Institutional versus public cases (table1)

While most studies utilized institutional samples to construct their datasets, two studies also incorporated public datasets (2, 16). One study relied solely on a public dataset (18). Tsou and Wu leveraged the large public TCGA thyroid cancer dataset, comprising H&E-stained whole slide images from PTC patients.

They obtained 51 and 52 samples with RAS and BRAF mutations, respectively, along with diagnostic labels, reducing the need for extensive in-house data collection and annotation (18). Dolezal et al. and Bohland et al. used both institutional and public cases. Bohland et al. evaluated two datasets - the Tharun dataset, aggregating cases from two centers to obtain 154 slides spanning 5 thyroid tumor subtypes, and the public 138-case Nikiforov collection (2). Dolezal et al. supplemented 115 in-house WSIs with 497 public TCGA cases (16). The remaining studies required the compilation of primary institutional datasets due to the lack of suitable public resources for their specific research questions. Sample sizes ranged from 25 institutional thyroid cancer cases (13 TERT-mutant, 12 TERT-wildtype) analyzed by Kim et al. (17) to over 1,300 institutional WSIs gathered by Zhu et al. (21).

All studies scanned images at either 20x or 40x magnification. While larger datasets enable more robust deep learning model development, they require greater resources for annotation and training.

Both institutional specimens and public datasets can provide representative histopathology data to develop and evaluate deep learning solutions. Overall, careful consideration of research aims to guide data preparation remains critical for continued progress in this emerging field.

### 4.4 Image acquisition technique (RGB vs Hyperspectral)

RGB was the standard image format across most studies. To improve model performance, some authors experimented with hyperspectral (HS) image acquisition. Two studies utilized HS images (22, 23), captured across a wide spectral range beyond human vision. Each pixel contains a continuous spectrum of measurements rather than just RGB values, enabling detailed spectroscopic histopathology analysis.

Tran et al. (23) designed a CNN model called HyperDeep for pixel-wise prediction of thyroid cancerous regions in HS images. HyperDeep achieved 95.3% average accuracy, outperforming benchmark CNNs using RGB images. Similarly, Halicek et al. (22) developed the Hyperspectral Tumor Detection Network (HTDN) for HS images, achieving over 93% tumor detection accuracy, surpassing conventional histopathology.

Overall, the combination of information-rich HS data and deep learning algorithms demonstrated promise for improving cancer diagnosis and evaluation.

### 4.5 Image preprocessing techniques

Many studies incorporated image preprocessing strategies to improve model performance. Kim et al. (17) experimented with color transformations like HSV (hue, saturation, value) and HED (hue, exposure, dark) in different combinations to enhance histological image details. The best performance was achieved using DenseNet161 (HSV-strong) + CRNN (HSV-strong). Wang et al. (3) used a Laplacian of Gaussian (LOG) filter to identify nuclei that appear dark under H&E staining.

### 4.6 Tumor segmentation/annotation (manual vs automated)

Both automated and manual approaches were utilized for tumor delineation. Automated techniques like CNN-based segmentation expedited the process, while manual annotation by pathologists ensured patches contained representative tumor regions. Kim et al. used CNN-based tumor detection and segmentation (17), while other studies relied on manual annotation. In Bohland et al. study (2), multiple pathologists provided diagnoses and manually annotated regions of interest (ROIs) on WSIs. Complexities arise from high-resolution WSIs, need for expert annotations, and variability in imaging conditions. However, diagnostic and slide-level labels in public datasets (16) enabled training neural networks without new annotations.

### 4.7 Extraction of patches/tiles (manual vs automated)

Cropping multiple patches from ROIs or entire WSIs was commonly used across studies to simplify analysis and expand limited datasets. However, patch-based approaches, whether manual or automated, may suffer from selection bias. Only two studies utilized manual patch extraction guided by pathologists(2, 24), while all other studies adopted automated extraction. Due to tumor heterogeneity, automated approaches require additional steps for tumor localization and annotation. Extracted patch sizes (table2) ranged from 2392 × 2392 pixels (20) down to 256 × 256 pixels (17), depending on magnification and neural network input requirements. Resizing large patches to match network defaults may lead to loss of fine details or cropping may discard relevant information, potentially impacting model performance. Conversely, very small patches could cause overfitting, as models may memorize patch-specific details rather than learning generalizable features applicable to the full dataset. Ultimately, patch size selection involves balancing trade-offs between capturing relevant information, computational constraints, and model requirements.

### 4.8 Classification approaches: DL vs traditional image analysis

Most studies relied on CNNs for classification of thyroid histopathology images, using state-of-the-art architectures like DenseNet, ResNet, VGG, and Inception. Pre-training on natural images (ImageNet) provides feature learning before fine-tuning on histology data, although the domain shift remains a consideration.

While DL models can learn discriminative features independently, providing engineered features as additional inputs may improve performance where labeled data is limited. Some works opted for traditional pipelines using hand-crafted features and ML classifiers, suggesting hybrid approaches could be beneficial (2). Bohland et al. compared traditional feature engineering to DL models. They performed nuclear segmentation with U-Net, extracted 36 nuclear features, and classified using ML. They aggregated median and standard deviation statistics across 700,000 nuclei per case to reduce features while preserving relevant information. Although slightly better performing, this approach requires extensive effort.

### 4.9 Base models (table2)

Most studies leveraged ImageNet-pretrained CNNs alone or combined with other components like LSTMs, MLPs, or traditional classifiers (table2). Model choices depend on factors like task complexity, compute resources, and performance needs.

Kim et al. (17) used DenseNet161, VGG16, and EfficientNet_b4 for detection, then a CRNN (ResNet152 + LSTM + MLP) for prediction. Halicek et al. (22) used InceptionV4 and MLP for detection. Some compared multiple CNNs like ResNet, DenseNet, VGG16, and EfficientNet (2, 3, 19, 24). Others used single models like Xception (16), InceptionV3 (18, 20), UNET (21), or VGG19 (23).

CNN model differences lie primarily in their base architectures, which vary in depth, efficiency, and design. Fine-tuning pretrained models or extending them with additional layers targets specific histopathology tasks. Recent works describe training CNNs from scratch on histopathology images rather than ImageNet (25).

### 4.10 Output class configuration (table 2)

Within the scope of the 11 scrutinized research endeavors, it is discernible that investigators meticulously tailored their classification tasks to harmonize with the clinical context and overarching objectives. This involved the utilization of both binary classifications, which accentuated pivotal diagnostic distinctions, and multi-class schemes. These strategic choices were influenced by the specific histopathology image datasets under examination and their corresponding clinical applications.

**Table 2:** Summary of the models’ architecture used across the studies with their performance metrics.

| Paper | Patch/tile size | Model Architecture | Output classes | AUC | Accuracy |
| --- | --- | --- | --- | --- | --- |
| Deng et al. 2023 | 2500×3200 pixel | ResNet50, ResNext50, EfficientNet, DenseNet121 | PTC, MTC, FTC, Adenomatous goiter, Adenoma, Normal | 0.822 - **0.994** | 92.18 - **97.53%** |
| Bohland et al. 2021 | 1916×1053 pixel<br>2272×2272 pixel | SVC (Feature-based)<br>ResNet101 (Deep learning) | PTC-like, non-PTC-like | 0.93<br>- | 89.7%<br>89.1% |
| Wang et al. 2019 | 448×448 pixel | VGG19<br>Inception-ResNetV2 | PTC, MTC, FTC, ATC, Goiter, Adenoma, Normal |  | **97.34%**<br>94.42% |
| Zhu et al. 2022 | 512×512 pixel | U-net, Decision tree | Tumor detection | 0.94 - **0.98** | - |
| Li et al. 2020 | 2392×2392 pixel | InceptionV3 | Benign, Uncertain, Malignant | - | < 0.8 (Patch level)<br>95.3% - 100 % (Slide level) |
| Chen et al. 2020 | 1024×1024 pixel | InceptionV3, VGG16, ResNet50 | Benign, Uncertain, Malignant | - | > 0.96 (ROI-based) |
| Kim et al. 2023 | 256×256 pixel | DenseNet161, VGG16, EfficientNet-B4 (detection)<br>ResNet152 (Feature extraction)<br>RNN (Feature integration)<br>MLP (TERT prediction) | TERT mutant<br>TERT wild-type | 0.94 | 0.95 |
| Dolezal et al. 2021 | 299×299 pixel | Xception-based | BRAF-like, RAS-like | 0.935 | - |
| Tsou & Wu 2019 | 2048×2048 pixel | InceptionV3 | BRAF, RAS | 0.94 | - |
| Tran et al. 2022 | 400×400 pixel | VGG19 | Tumor detection | 0.96 (HS)<br>0.93 (RGB)<br>0.94 (HS-synthesized RGB) | - |
| Halicek et al. 2020 | 1040×1392 pixel | InceptionV4, MLP | Tumor detection | 0.9 (HSI-synthesized RGB) | - |

A predominant tendency among these studies was to approach histopathology image classification primarily through the lens of binary classification. This binary paradigm was often adopted for purposes such as subtyping, mutation prediction, or tumor segmentation. For instance, Bohland et al. (2) embarked on a subtyping endeavor, categorizing thyroid nodules into either PTC-like or non-PTC-like. In the domain of mutation prediction, Kim et al. (17) ascertained the TERT promoter mutation status, classifying it as either positive or negative, while Tsou et al. (18) endeavored to differentiate between RAS and BRAF mutated subtypes. Similarly, Dolezal et al. (16) dichotomized thyroid nodules into RAS-like or BRAF-like mutated subtypes. In the context of tumor segmentation, scholars such as Tran et al., Halicek et al., and Zhu et al. (21-23) employed binary classification to segregate patches into tumor or normal categories.

Conversely, several studies departed from this binary dichotomy, opting for multi-class classification paradigms. Notably, Li et al. and Chen et al. (19, 20) extended their classifications to three categories: benign, malignant, or uncertain. Wang et al. (3) pushed the envelope further by defining a comprehensive taxonomy of seven classes, encompassing normal, goiter, FA, PTC, FTC, MTC (medullary thyroid carcinoma), and ATC (anaplastic thyroid carcinoma). In a similar vein, Deng et al. (24) orchestrated the classification of thyroid lesions into one of six classes: PTC, MTC, FTC, FA, adenomatous goiter, or normal.

The choice between binary and multi-class classification hinges on a confluence of factors, including the clinical inquiry at hand and the inherent characteristics of the data. In navigating this decision-making process, it is imperative to deliberate upon the clinical significance of model predictions and the potential ramifications thereof. For instance, in scenarios where the diagnostic course of action diverges significantly between subtypes, a multi-class approach emerges as a more judicious choice. Multi-class classification affords a finer level of granularity in predictions, catering to the nuances of distinct pathological categories. Conversely, when the clinical decision ultimately resolves into a binary outcome, especially within the framework of an imbalanced dataset, the adoption of a binary-output model is preferred, given its simplicity and interpretability. Careful consideration of these factors is paramount in ensuring that the chosen classification strategy aligns harmoniously with the clinical context and objectives.

### 4.11 Analysis of Results/Evaluation of model performance (Table2)

The studies consistently demonstrated the potential of deep learning to extract discriminative features from thyroid histology images, enabling accurate classification and prediction. Studies utilized various evaluation metrics to assess model performance on image classification tasks. Accuracy and AUC (area under the ROC curve) were most common, measuring overall correctness of predictions and ability to discriminate classes, respectively.

Reported accuracy ranged from 77-100%, with substantial variability attributable to differences in study design, methods, and datasets. However, most studies achieved high accuracy of 90% or above, with top-performing models reaching 80-95% accuracy.

Notably, Kim et al. (17) achieved 0.95 accuracy and 0.94 AUC using color transformation in preprocessing. Bohland et al. (2) reported DL accuracy of 77.4% and 89.1% on two datasets, versus 83.5% and 89.7% for features-based, with mean AUC of 0.93 and 0.75. Multiple studies achieved AUC exceeding 0.9 (16, 18, 22, 23) (Fig. 2).

**Figure 2:**
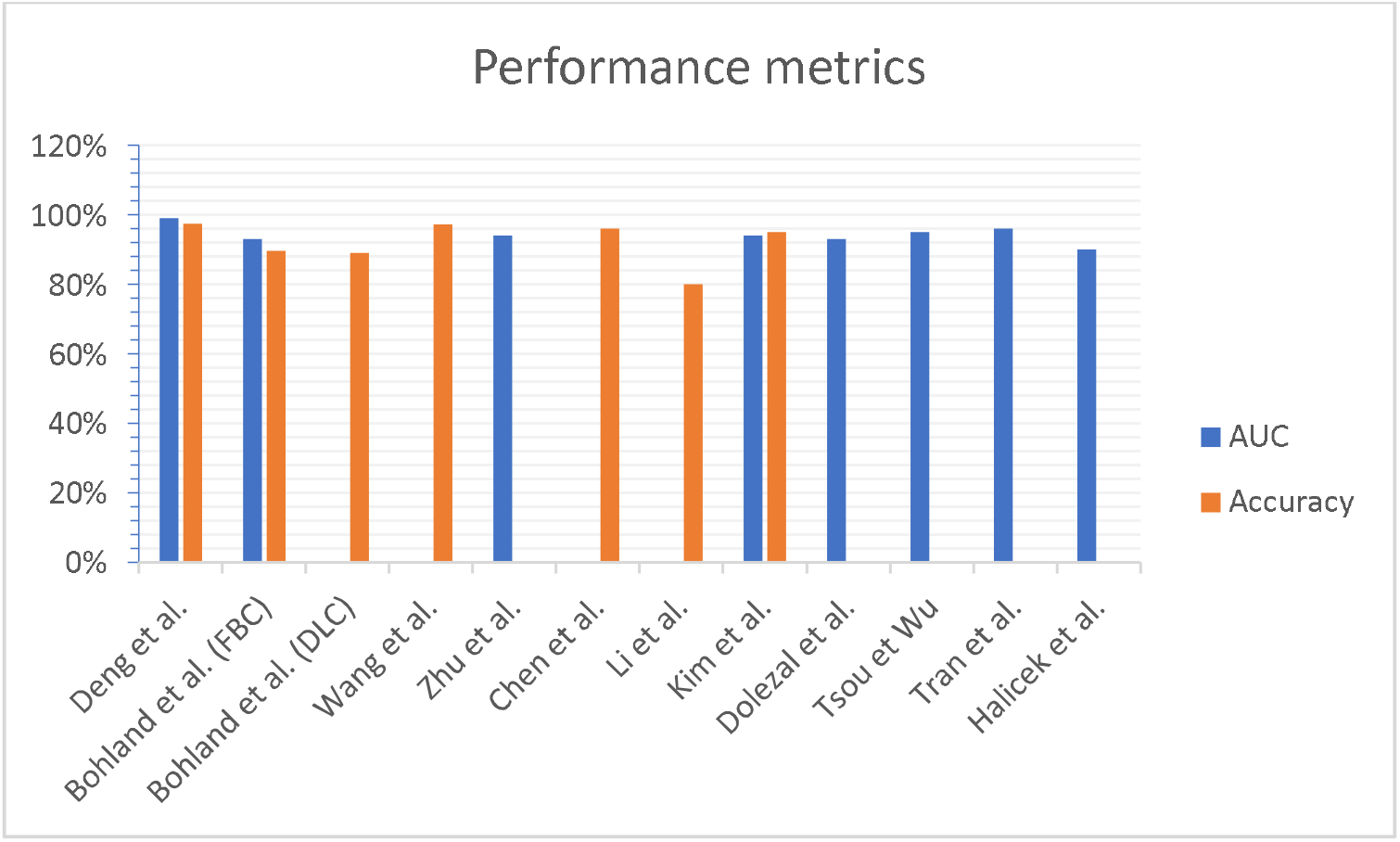
Comparison of performance metrics among the studies

Some authors reported two levels of performance assessment. Li et al. (20), reported patch-level accuracy of <80% and slide-level accuracy (with rule-based method) of 95.3-100%. Similarly, Chen et al. (19) reported a classification accuracy of > 0.96 on ROI-based diagnosis.

While these results offer promising insights into the potential of DL in advancing thyroid diagnosis and management, further research is warranted to address identified limitations.

## 5. Limitations with suggested key solutions

The selected studies demonstrated promising applications of DL for thyroid histopathology image classification. However, some key limitations were observed that restrict the real-world utility and clinical translation of these approaches. Small sample sizes, subjective annotation, and lack of case diversity were common limitations. The works highlight the need for larger standardized datasets, expanded diversity in subclasses, comparison across multiple sites and rigorous evaluation on heterogeneous real-world data.

### 5.1 Small, imbalanced dataset

One fundamental challenge is the small, imbalanced datasets used in most studies, which limits model performance and generalizability (16). Furthermore, diversity is limited given single-center data (17, 23). DL prerequisites of large, labeled datasets and computational resources may hinder its translation to clinical practice. Expanding datasets through extensive labeling efforts with increased examples per class will be critical moving forward.

### 5.2 No quality control for data preparation

Objective quality control of data extraction and preprocessing programs is required to limit potential biases (21). Direct comparison of pathologists with versus without assistance of models using reader studies would better demonstrate clinical utility and guide development. There is also a lack of head-to-head benchmarking between techniques on shared public datasets (2).

### 5.3 Bias in ROI annotation and patch extraction

Additionally, potential subjectivity and selection bias can arise during tumor/region annotation by pathologists, which may not fully capture the diversity of real-world samples. Also, subjective patch sampling may introduce selection bias. Developing standardized annotation protocols could help mitigate this (17). Most studies focused narrowly on classification tasks, while localization, segmentation, and other problem formulations warrant deeper investigation.

### 5.4 lacking details for model deployments

As an emerging field, translating these approaches into widespread clinical practice remains challenging. Crucially, clinical implementation remains overlooked, with no clear strategies for model deployment and workflow integration. Conducting usability studies is advised before deployment.

### 5.5 Model optimization & interpretability

Regarding model development, the field is still in early stages, with limited model architectures and hyperparameters evaluated so far. Broad optimization of network architectures and hyperparameters through extensive experiments could boost performance. Another key aspect is model interpretability, which remains underexplored but critical for clinical acceptance. Implementing methods to explain model reasoning and validate learned features is beneficial.

### 5.6 Lacking clinical and radiological correlation

Another issue is the sole reliance on histopathology images, lacking complementary clinical, genomic, and radiographic data that pathologists use to reach diagnostic decisions. Integrating these multiple modalities represents an important frontier. Enabling multi-modal integrations represent fertile directions for future research to unlock the full potential of DL in thyroid histopathology.

## 6. Future Directions

The findings reveal the potential of DL approaches in enhancing the accuracy of thyroid neoplasms classification. Integrating DL methods into daily practice looks optimistic in enhancing thyroid neoplasms diagnosis. The high accuracy observed in these studies suggests their potential utility in aiding clinical decision-making. Initial results, though limited in scale, strongly demonstrate the capability to encode morphological patterns in an automated high-throughput manner. This could enhance pathologist productivity and improve patient care through quantitative augmented decision support.

Looking ahead, larger multi-center studies, evaluation on heterogeneous real-world data, and pragmatic clinical integration will be crucial to fully translate these emerging technologies into widespread practice. However, the early successes establish the way for larger initiatives to unite pathologists, data scientists and clinicians in realizing the next generation of AI-powered computational thyroid pathology.

## 7. Conclusion

This systematic review offers comprehensive insights into recent advancements in DL approaches for the classification of thyroid neoplasms based on histopathology images. The findings underscore the potential of these techniques to enhance accuracy and improve clinical decision-making. While further optimization is warranted, integration of these tools to guide pathologists could markedly improve thyroid cancer screening, diagnosis, prognosis, and treatment selection for better patient care.

## Data Availability

All data produced in the present work are contained in the manuscript

## Declaration of generative AI and AI-assisted technologies in the writing process

During the preparation of this work the authors used ChatGPT and google Bard only in order to improve readability and language. After using this service, the authors reviewed and edited the content as needed and take full responsibility for the content of the publication.

